# EEG responses to auditory stimuli are less context-dependent in preschoolers with autism spectrum disorder compared to typical development

**DOI:** 10.64898/2026.04.17.26350631

**Authors:** Meiwen Shao, Katie A. McNair, Gerardo Parra, Celia Tam, Nancy Sullivan, Damla Şentürk, Jeffrey P. Gavornik, April R. Levin

## Abstract

Individuals with autism spectrum disorder (ASD) often exhibit atypical auditory processing, yet it remains unclear whether and how the integration of simple acoustic features and contextual information is impacted in ASD. One real-world example of this integration is the auditory looming bias, the prioritized processing and perception of approaching auditory stimuli. We designed a paradigm that presents intensity-rising (looming) and intensity-falling (receding) auditory stimuli to 3–4-year-old children with ASD (n = 21), children with sensory processing concerns who do not have ASD (SPC; n = 16) and children with typical development (TD; n = 30). We recorded neural responses using electroencephalography (EEG) and found evidence of looming bias in the SPC and TD groups, as indexed by greater P1 peak amplitude during the looming than receding stimuli (TD: t(64) = 6.87, p < .001; SPC: t(64) = 4.07, p < .001). But this finding was not present in the ASD group (p = .194). Additionally, the ASD group showed reduced differentiation between looming and receding stimuli, as indicated by significantly lower Rise-Fall Difference Score (RFDS) in comparison to the TD group (Z = −3.00, *p_adj_* = .008). These findings suggested altered context-dependent modulation of sensory input in ASD.

**Lay Summary:** Many children with autism show differences in how they process sounds. Using sound patterns in which loudness gradually increased and decreased over time, like many real-world sounds, we found that children with autism showed less neural differentiation between increasing and decreasing sounds. This finding suggested that the brain may process changes in sound differently in autism, particularly in how it adjusts to sounds as they change over time, which could contribute to the sensory challenges many children with autism experience in daily life.

## Introduction

### Autism and Sensory Processing

Autism spectrum disorder (ASD) is a neurodevelopmental disorder characterized by clinically and functionally significant deficits in two primary domains: social communication and restricted, repetitive behaviors (1). Atypical sensory processing is one of the core features of restricted, repetitive behaviors associated with ASD diagnosis and can occur in a range of sensory modalities (2), including auditory, visual, tactile, gustatory, olfactory, and interoception. Notably, hyper- and hyporesponsivity often co-occur within the same individual, a phenomenon known as the Sensory Paradox (3). For example, one may cover their ears to vacuum cleaner noise (hyperresponsivity) but not respond to name-calls (hyporesponsivity). Studying how the interplay between contextual factors (e.g., the temporal dynamics of stimuli) and basic stimulus features (e.g., intensity or salience) influence sensory processing may help reveal the mechanisms underlying this paradox.

Although individuals with ASD often exhibit atypical responses to sound, basic auditory function is typically preserved, and an associated risk of hearing loss for individuals with ASD has not been supported (4). These differences are instead thought to reflect changes in neural circuitry (5,6), leading to altered neural encoding and perception of sensory stimuli that can eventually interfere with the typical development of higher-level behaviors such as social communication, daily living skills, and community participation (7,8).

Importantly, atypical sensory processing is not exclusive to ASD (9). Atypical sensory processing has been found to be present in many other neurodevelopmental and psychiatric disorders such as attention-deficit/hyperactivity disorder, specific learning disorder, and depressive disorders (10). Furthermore, atypical sensory processing can also be present in individuals who do not meet criteria for any neurodevelopmental or mental disorders (11). It is an ongoing scholarly discussion whether clinically significant atypical sensory processing (often called “sensory processing disorder”) should be considered a standalone diagnostic entity or a non-specific transdiagnostic phenotype to neurodevelopmental or neuropsychiatric disorders. For this investigation, the term sensory processing concerns (SPC) is used to describe individuals with caregiver-reported sensory concerns but do not meet criteria for an ASD diagnosis.

Understanding how sensory symptoms manifest differently in individuals with ASD, SPC, and Typical Development (TD) is important to identifying ASD-specific biomarkers and validating (or invalidating) sensory processing disorder as a standalone diagnosis.

### Auditory Looming Bias

Looming auditory stimuli, which are characterized by dynamically rising intensity, are usually associated with approaching objects or attention-capturing cues. For example, calling one’s name in increasing intensity is a way to get their attention. The tendency to encode looming stimuli as salient and thus to allocate more attentional and physiological resources to their perception is called the looming bias. Investigating looming bias in a laboratory setting allows for the examination of the contextual perception of sounds. In comparison to experimental paradigms that test one dimension of the stimulus (e.g., loudness or a single temporal process such as habituation) in isolation from other dimensions, testing looming bias presents greater ecological validity because separate dimensions of a stimulus are not isolated in real-life.

Previous behavioral studies in healthy adults have shown evidence of looming bias: when compared with receding or static sounds, looming sounds have been associated with shorter estimation of time to arrival (12), greater perceived loudness change (13), and faster perceived moving speed (14). In addition to behavioral studies, neurophysiological studies have also found evidence of looming bias. A human magnetoencephalography (MEG) study showed that the absolute field strength for a complex (i.e., non white-noise) sound with rising intensity is greater than that of the same sound with falling intensity (15). Human functional magnetic resonance imaging studies have also shown evidence of the looming bias in the form of greater blood oxygen level-dependent responses (16,17). Electroencephalography (EEG) studies have identified looming bias in event-related potentials (ERPs), with notable effects in the amplitude of the P2 component (18), the latency of a negative component measured at the prefrontal cortex (19), and the amplitude of the N1 component (20).

Although auditory looming bias has not, to our knowledge, been examined in ASD, prior research has characterized neural alterations in temporal aspects of auditory processing in this population. Two canonical EEG paradigms in the field are the oddball paradigm and the paired-click paradigm, which investigate auditory change-detection and sensory gating, respectively. Studies using the oddball paradigm have found evidence of greater (21,22) and reduced (23) mismatch negativity (MMN) amplitude in children with ASD, suggesting that the salience of an auditory oddball may be bidirectionally altered in ASD relative to healthy controls, with directionality likely influenced by stimulus properties, the listener’s developmental stage, and auditory sensitivity subtype. Paired-click studies show evidence of P50 (24,25) and N2 (26) gating deficit in children with ASD, suggesting reduced use of a prior stimulus to modulate response to a target stimulus relative to healthy controls.

### The Present Study

In this study, we extended prior work on isolated auditory temporal processes by presenting intensity-rising (looming) and intensity-falling (receding) stimuli. Because the perception of looming and receding sounds recruits several temporal processes—such as detecting changes, contextualizing repeated signals, and predicting upcoming stimuli—this approach allows multiple temporal mechanisms to be examined within a single paradigm in a way that better reflects real-world auditory experience. Additionally, intensity-defined looming and receding stimuli inherently allow us to examine how intensity interacts with temporal context, clarifying how multiple dimensions of auditory processing jointly shape the perception of sounds. We used EEG to measure brain activity in ASD, SPC, and TD children, 3–4 years of age, as they listened to auditory stimuli with an intensity-rising and an intensity-falling phase. We particularly target children in early development because prior work has shown that atypical sensory processing in ASD emerges early in life (27) and can shape downstream developmental trajectories (7). First, we aimed to assess response during the intensity-rising phase compared to the intensity-falling phase of the auditory stimuli, in each group separately. We hypothesized that the TD group would show greater neurophysiological response to the intensity-rising than intensity-falling phase, indicating the presence of looming bias. We then aimed to compare looming responses across groups. We hypothesized that the ASD and SPC groups would show less looming bias than the TD group, pointing to less efficient use of context to modulate neural responses to auditory stimuli.

## Materials and Methods

### Participants

Sixty-seven 3-to 4-year-old children (21 ASD, 16 SPC, and 30 TD) participated in the present study. Participants in the ASD group had community diagnoses of ASD and met criteria for ASD based on information obtained during the study visit. All participants in the SPC group had parent or caregiver concerns about how the child processed sensory information but had not received an ASD diagnosis and did not meet the criteria for ASD based on information obtained during the study visit. To include a broad array of ASD subtypes and potential circuit-level abnormalities, participants were not excluded from the ASD or SPC groups for having known risk factors for neurodevelopmental conditions (e.g., prematurity, neurogenetic disorders) or neuropsychiatric comorbidities. TD participants had no known or suspected ASD diagnosis, and no other known or suspected neurodevelopmental or neuropsychiatric disorders. All participants reported normal hearing, no physical impairments that would preclude participation in testing, no regular use of medications known to affect sleep or cognition, and English was spoken at home more than half of the time.

Several sources of information were used to confirm group placement during the study visit. All participants in the ASD group completed the Autism Diagnostic Observation Schedule – 2^nd^ edition (ADOS-2; (28)). In the SPC group, the ADOS-2 (14 of 16 participants) and/or the Social Communication Questionnaire (SCQ; (29)) (16 of 16 participants) were administered to confirm that there was no concern for autism. All TD participants also obtained group confirmation through the SCQ. Because the sensitivity and specificity of ADOS-2 for ASD are approximately 90% and 83% respectively (30) (i.e., it is expected that some individuals will meet ASD criteria on the ADOS-2 but not have ASD, and vice versa), additional clinical information (including medical record review and/or diagnostic interview) was obtained in any participant for whom group placement reported by the parent did not match ADOS-2 findings. For example, this applied to cases in which participants had parents who reported that they should be in the SPC group but classified as autism spectrum disorder or autism using the ADOS-2, and participants whose parents reported they had received a prior ASD diagnosis but did not end up meeting criteria on the ADOS-2. When correct group status could not be determined based on information obtained through the study visit, a behavioral neurologist specializing in ASD (ARL) saw the patient in clinic through the Autism Spectrum Center at Boston Children’s Hospital to determine group status. Additionally, since a diagnosis of ASD requires that challenges with social skills are not better accounted for by cognitive delay or intellectual disability, all participants meeting criteria for ASD on the ADOS-2 had testing with the Mullen Scales of Early Learning (MSEL; (31)) to confirm a nonverbal mental age of at least 18 months. Nonverbal mental age was calculated by averaging the age equivalent of the visual reception scale and the fine motor scale. Three participants whose parents initially reported that they should be in the ASD group were excluded from the study after a nonverbal mental age of less than 18 months was found on the MSEL; these three were not included in the total n = 21 of the ASD group. One ASD participant did not complete the MSEL during the study visit but was later confirmed via medical record review to have a nonverbal mental age of ≥ 18 months and is therefore included in the n = 21.

Participants were recruited from local clinics, the Research Participant Registry through the Translational Neuroscience Center at Boston Children’s Hospital, community flyers, and community outreach events. Participants in the ASD group were also recruited using the Simons Foundation Powering Autism Research for Knowledge (SPARK; (32)) database. The study was approved by the Institutional Review Board at Boston Children’s Hospital.

### Experimental Stimuli

Participants were presented with 50 cycles of 14 250 ms auditory tones, with tone loudness following a step function that increased from 53 to 70 dB SPL (53, 59, 63, 65, 67, 69, 70 dB SPL) and then decreased through the same levels (Fig. 1). Tones were created, delivered, and synced with EEG recording through the Presentation software (Neurobehavioral Systems, Inc., Berkley, CA, USA). Each tone was created by multiplying a 500Hz carrier sine wave with a 32Hz amplitude modulation sine wave. To reach the tone’s desired loudness, its waveform was further multiplied by a constant. Tones were presented continuously within each cycle, with a 500 ms silent interval between cycles. All tones were presented using Logitech Z150 Stereo Speakers, and the total duration of the paradigm was 3 minutes and 20 seconds.

**Fig. 1.**
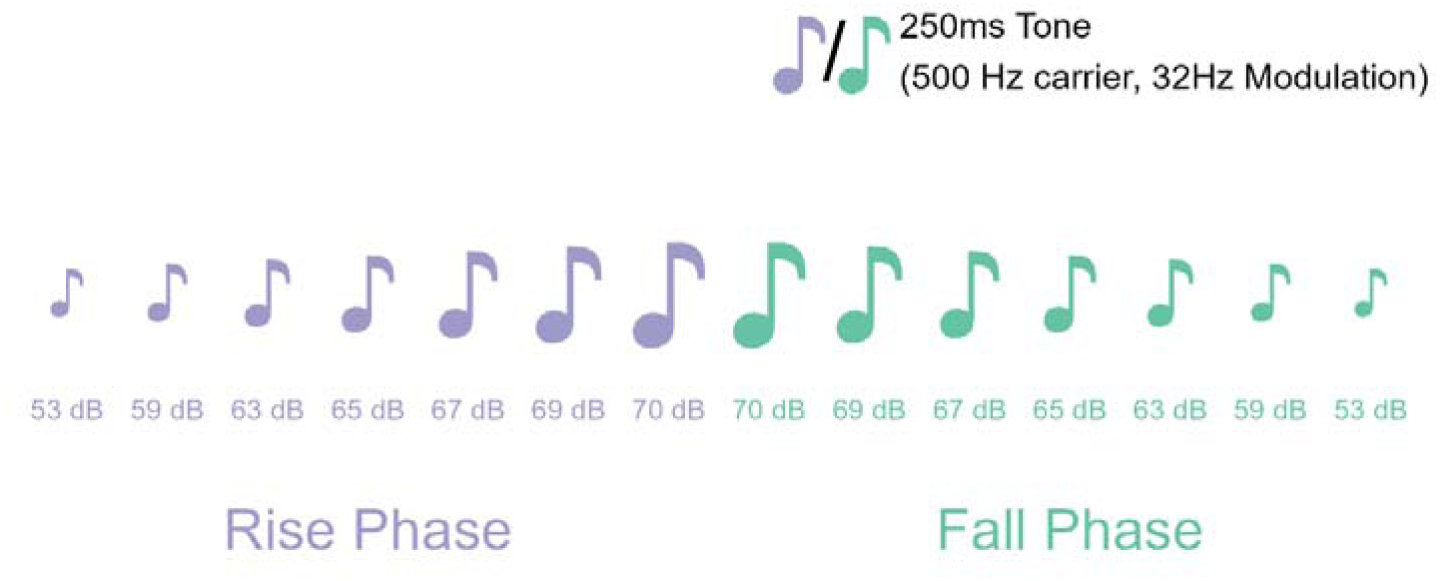
Experimental schematic illustrating one tone cycle. The depicted cycle is presented 50 times per experiment, with a 500 ms silent interval between cycles.

### Procedure

Prior to the study visit, families received via email a video-based social story to review, which explained the study procedures and helped children and families prepare for the study visit. On visit day, participants were seated in a minimally lit, electro-acoustically shielded booth next to a behavioral assistant who provided brief corrective instructions (e.g., “Stay still” or “Keep quiet”) as needed to optimize recording quality. For participants who needed more behavioral support, a parent or caregiver sat beside the participant or held the participant on their lap. Participants watched the movie WALL-E (33) on silent while the auditory stimuli were presented. The use of a standardized silent movie to enhance compliance is standard in ERP studies of neurodevelopmental disorders and in this age group (34).

### EEG Acquisition and Processing

EEG data were collected with 128-channel Hydrocel Geodesic Sensor Nets with NetAmps 400 amplifier through the NetStation software (Magstim EGI, Eugene, OR, USA). The electrooculography channels (i.e., 125, 126, 127, 128) were removed prior to recording to prioritize participant comfort. EEG data were digitized at 1000Hz and referenced online to the vertex (i.e., electrode Cz).

EEG Data were preprocessed with the Harvard Automated Processing Pipeline for Electroencephalography plus Event-Related (HAPPE+ER) version 4 (35,36) in MATLAB R2023a (Mathworks, Inc.). First, forty-four channels were selected for pre-processing (Fig. 2), providing coverage across all major scalp regions. Next, the electrical line noise at 60Hz was removed using CleanLine (37). Then, a bandpass Butterworth highpass filter of 0.1Hz and lowpass filter of 30Hz were used and bad channel rejection took place (see Section 2.6 in (35)). Data were then processed with soft wavelet thresholding before segmentation, during which data were segmented into epochs extending from 100 ms prior to and 249 ms after the onset of each tone. The epochs were then baseline corrected using the −50 ms to 0 ms pre-stimulus window, and bad data interpolation was performed, such that data determined to be artifact-contaminated within each epoch was rejected and subsequently interpolated with spherical splines.

**Fig. 2.**
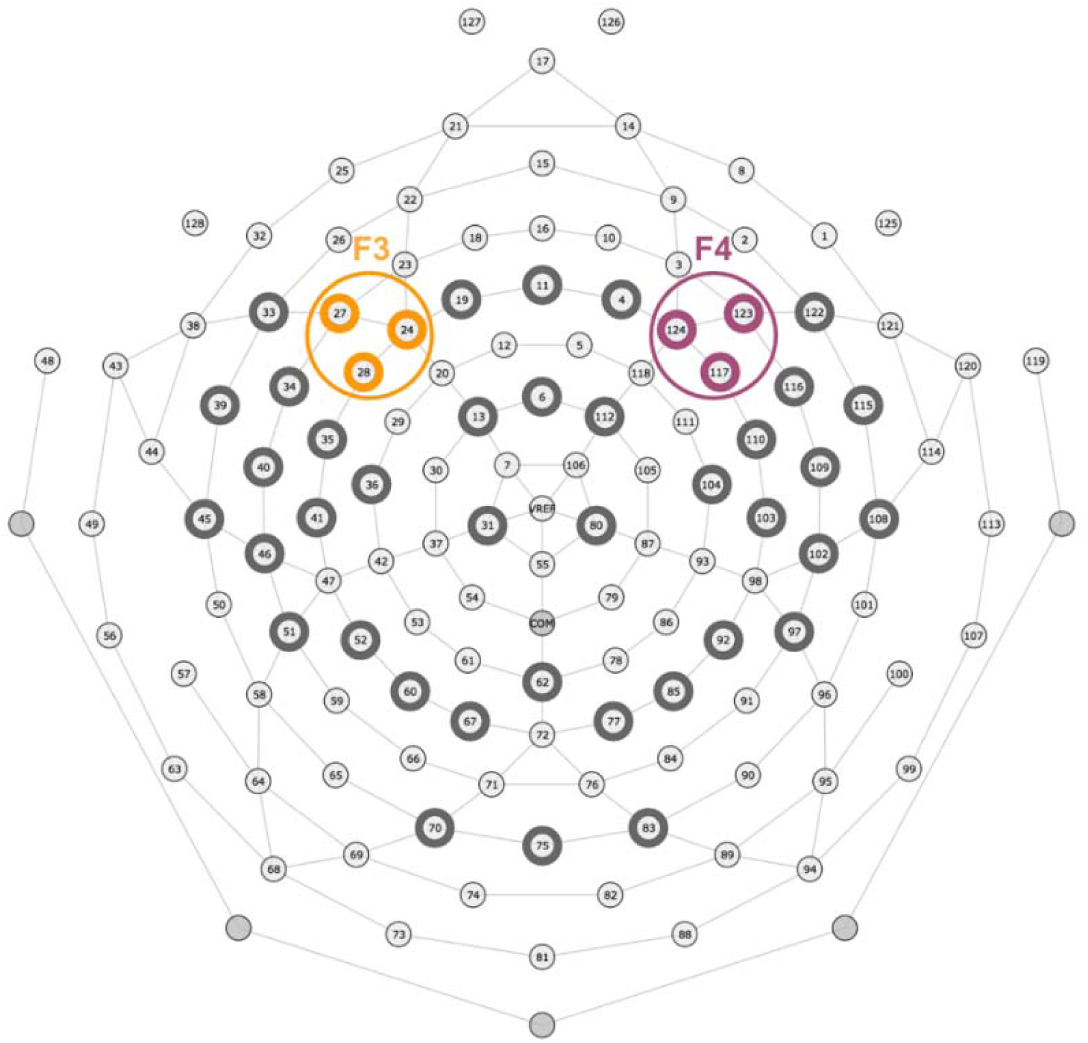
44 channels entered EEG data preprocessing (channels with thick outlines), out of which 6 were selected for data analysis – 3 channels belonged to the F3 cluster (orange), and 3 channels belonged to the F4 cluster (magenta).

Any epoch in which voltage at any channel at any time exceeded 150 μV or was less than −150 μV was rejected. Then, channels removed during the bad channel rejection step were subjected to spherical interpolation. Lastly, data were re-referenced to average reference.

### Event-Related Potential (ERP) Processing

We conducted ERP analyses in two stages. First, we examined overall differences between responses during the rise (looming) and the fall (receding) phase of the auditory stimuli. To this end, we extracted our ERP time-series of interest by averaging voltage across a cluster of 6 channels (Fig. 2) centered on the F3 and F4 channels. We defined the first 7 tones in the tone cycle (53, 59, 63, 65, 67, 69, 70 dB SPL) as the rise phase of the cycle, and the remaining 7 tones (70, 69, 67, 65, 63, 59, 53 dB SPL) as the fall phase of the cycle (Fig. 1). To obtain the average rise phase ERP time-series for each participant, we calculated the mean ERP time-series of all trials in all rise phase tones and then repeated this step to obtain the fall phase average ERP time-series (Fig. 3A-C).

**Fig. 3.**
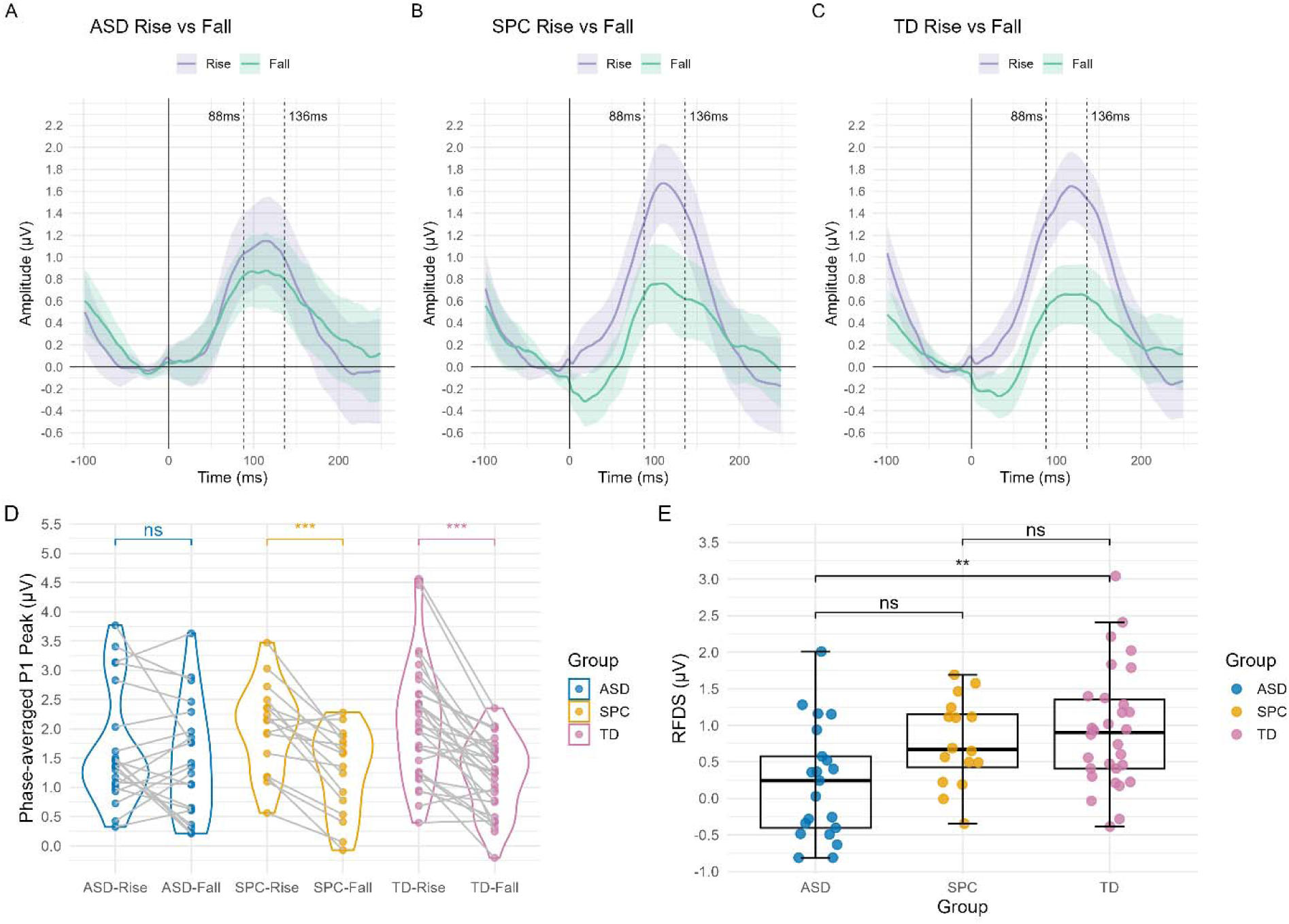
**A-C)** The rise phase and fall phase average voltage amplitudes extracted across a cluster of 6 electrodes centered on F3 (electrodes 24, 27, 28) and F4 (electrodes 117, 123, 124) in the ASD, SPC, and TD groups respectively. Shaded areas depict the standard errors of the means. **D)** SPC and TD groups show significant difference in the phase-averaged P1 peak amplitude (i.e., maximum peak amplitude 88-136 ms post stimulus onset) between the rise and fall phase. This difference was not seen in the ASD group. **E)** ASD and TD groups show significantly different Rise Fall Difference Score (RFDS). Significant RFDS difference was not seen in any other group comparisons. Levels of statistical significance: ns = not significant, \**p*<.05, \*\**p*<.01, \*\*\**p*<.001

Second, we studied whether looming bias reflects a consistent change in responses across all intensity levels or instead varies as a function of intensity. We examined responses separately at each intensity level within the rise and fall phases. Given prior work demonstrating that the P1 component reflects early auditory processing (38), we extracted the P1 peak amplitude for each stimulus as the absolute maximum amplitude 88 ms–136 ms post stimulus onset. The 88 ms–136 ms P1 time window was determined by a priori expectations (39–41) and adjusted based on the latency of the prominent peak in the grand-average Global Field Power (GFP) computed from the present study’s dataset, collapsed across all participants and trials.

### Statistical Analysis

For each participant, we obtained 14 P1 peak amplitudes, one for each condition, which were then analyzed in a mixed-design ANOVA with phase (i.e., rise vs fall) and stimulus intensity (i.e., the 7 intensities ranging from 53dB to 70dB) as within-subjects factors and group (i.e., ASD, SPC, or TD) as a between-subjects factor through the afex package (42) in R software (R Core Team, Vienna, Austria). Normality of residuals was verified by visual inspection of Q–Q plot; homogeneity of variances for between-subjects factor as well as sphericity for within-subjects factors were confirmed using the performance package (43) in R. Follow-up simple-effect analyses were conducted to examine the effect of phase within each group by using the emmeans package (44). To examine whether the size of the phase effect differed between groups, we computed average P1 peak amplitudes for the rise phase and fall phase separately. Subsequently, the Rise-Fall Difference Score (RFDS), calculated as:

RFDS (µV) = [Rise phase average P1 Peak] (µV) – [Fall phase average P1 Peak] (µV) was obtained for each participant. Group differences in RFDS data were then analyzed with a Kruskal-Wallis Test featuring group as a between-subjects factor, as ANOVA residuals were not normally distributed. Post-hoc Dunn tests were used to conduct follow-up comparisons. Dunn tests were performed with the FSA package (45) in R software, and p-values were adjusted with the Holm method.

## Results

Initial examination of group-and-phase-averaged ERP responses showed clear P1 peaks during the selected P1 window of 88 ms–136 ms (Fig. 3A-C) in all three groups and both phases, which demonstrated integrity in our selected EEG measure and motivated the subsequent analyses.

The ANOVA (Table 2) revealed a significant effect of phase, F(1, 64) = 45.61, p < .001, η²p = .416, and intensity, F(6, 384) = 2.29, p = .035, η²p = .035, on P1 peak amplitudes (i.e., maximum peak amplitude 88 ms–136 ms post stimulus onset). Additionally, we observed a significant interaction between group and phase, F(2, 64) = 5.94, p = .004, η²p = .157. Follow-up tests revealed a significant effect of phase on P1 peak amplitudes (Table 3, Fig. 3D) in the TD, t(64) = 6.87, p < .001, and SPC, t(64) = 4.07, p < .001, groups, but not the ASD group (p = .194). In both the TD and SPC groups, P1 peak amplitudes were larger in the rise phase (TD: M = 2.13, SD = 1.69, SPC: M = 2.02, SD = 1.48) than the fall phase (TD: M = 1.19, SD = 1.49, SPC: M = 1.26, SD = 1.43).

**Table 1.**
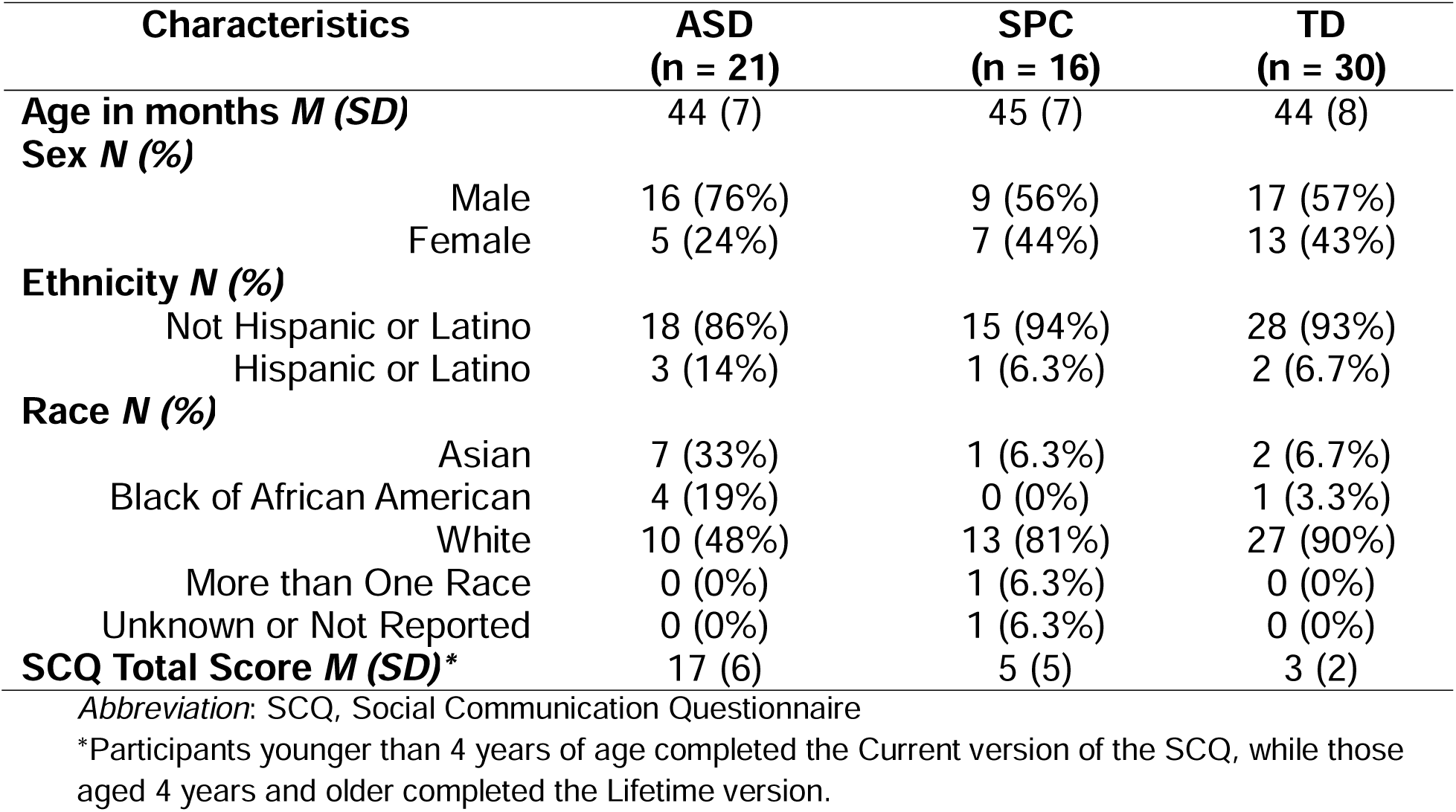
Participant Characteristics.

| **Characteristics** | **ASD**<br>**(n = 21)** | **SPC**<br>**(n = 16)** | **TD**<br>**(n = 30)** |
| --- | --- | --- | --- |
| **Age in months *M* (SD)** | 44 (7) | 45 (7) | 44 (8) |
| **Sex *N* (%)** |  |  |  |
| Male | 16 (76%) | 9 (56%) | 17 (57%) |
| Female | 5 (24%) | 7 (44%) | 13 (43%) |
| **Ethnicity *N* (%)** |  |  |  |
| Not Hispanic or Latino | 18 (86%) | 15 (94%) | 28 (93%) |
| Hispanic or Latino | 3 (14%) | 1 (6.3%) | 2 (6.7%) |
| **Race *N* (%)** |  |  |  |
| Asian | 7 (33%) | 1 (6.3%) | 2 (6.7%) |
| Black or African American | 4 (19%) | 0 (0%) | 1 (3.3%) |
| White | 10 (48%) | 13 (81%) | 27 (90%) |
| More than One Race | 0 (0%) | 1 (6.3%) | 0 (0%) |
| Unknown or Not Reported | 0 (0%) | 1 (6.3%) | 0 (0%) |
| **SCQ Total Score *M* (SD)\*** | 17 (6) | 5 (5) | 3 (2) |

*Abbreviation:* SCQ, Social Communication Questionnaire
\*Participants younger than 4 years of age completed the Current version of the SCQ, while those aged 4 years and older completed the Lifetime version.

**Table 2.**
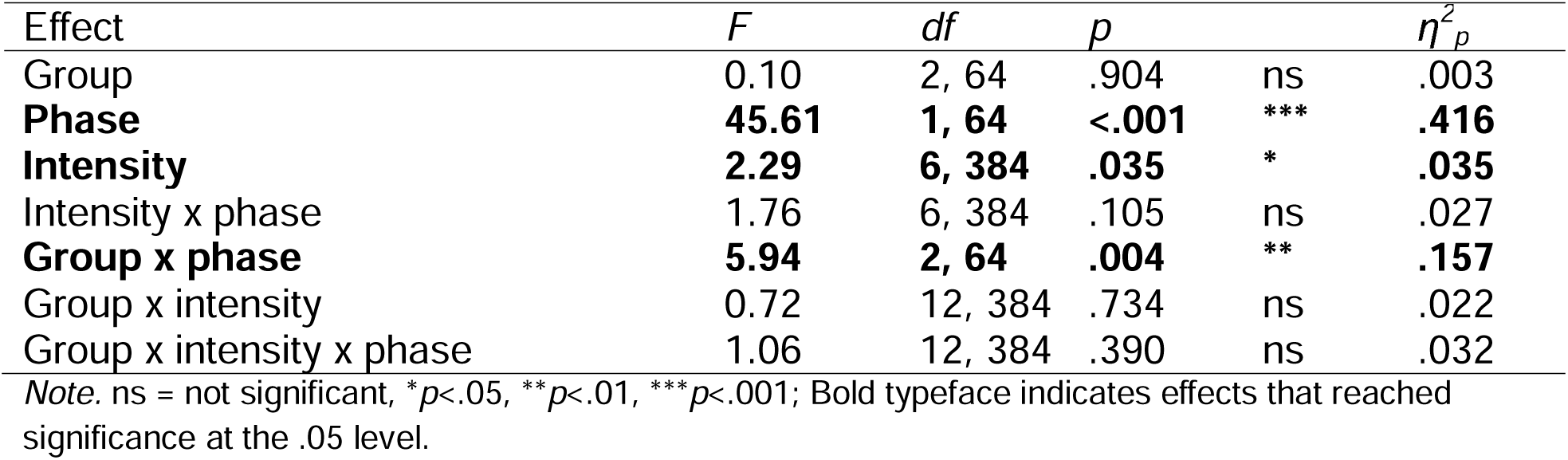
Mixed-Design ANOVA for P1 Peak.

| Effect | *F* | *df* | *p* | | $\eta^2_p$ |
| --- | --- | --- | --- | --- | --- |
| Group | 0.10 | 2, 64 | .904 | ns | .003 |
| **Phase** | **45.61** | **1, 64** | **<.001** | **\*\*\*** | **.416** |
| **Intensity** | **2.29** | **6, 384** | **.035** | **\*** | **.035** |
| Intensity x phase | 1.76 | 6, 384 | .105 | ns | .027 |
| **Group x phase** | **5.94** | **2, 64** | **.004** | **\*\*** | **.157** |
| Group x intensity | 0.72 | 12, 384 | .734 | ns | .022 |
| Group x intensity x phase | 1.06 | 12, 384 | .390 | ns | .032 |

Note. ns = not significant, \* $p < .05$ , \*\* $p < .01$ , \*\*\* $p < .001$ ; Bold typeface indicates effects that reached significance at the .05 level.

**Table 3.** Effect of Phase on P1 Peak Amplitude Within Each Group.

|  | EMM of Rise – Fall<br>P1 | SE | *df* | *t* | *p* |  |
| --- | --- | --- | --- | --- | --- | --- |
| ASD | 0.22 | 0.16 | 64 | 1.31 | .194 | ns |
| **SPC** | **0.77** | **0.19** | **64** | **4.07** | **<.001** | **\*\*\*** |
| **TD** | **0.90** | **0.14** | **64** | **6.87** | **<.001** | **\*\*\*** |

Note. EMM: estimated marginal mean; ASD: autism spectrum disorder; SPC: sensory processing concern; TD: typical development; ns = not significant, \* $p < .05$ , \*\* $p < .01$ , \*\*\* $p < .001$ ; Bold typeface indicates effects that reached significance at the .05 level.

Results of the Kruskal-Wallis test revealed a significant difference in RFDS between groups, x^2^(2) = 9.65, *p* = .008. The follow-up Dunn test (Table 4, Fig. 3E) showed a significant difference in RFDS between the ASD and TD group (Z = −3.00, *p_adj_* = .008), but not between the ASD and SPC group (p = .054), or between the SPC and TD group (p = .700).

**Table 4.** Kruskal-Wallis and Dunn Test results for RFDS.

Kruskal-Wallis and Dunn Test results for RFDS
|  | Comparison | Statistic | *P* (unadjusted) | *P* (adjusted) |  |
| --- | --- | --- | --- | --- | --- |
| **RFDS** | **Group** | $\chi^2 = 9.65$ | **0.008** | **NA** | **\*\*** |
| | **ASD-TD** | $z = -3.00$ | **0.003** | **0.008** | **\*\*** |
| | ASD-SPC | $z = -2.21$ | 0.027 | 0.054 | ns |
| | SPC-TD | $z = -0.39$ | 0.700 | 0.700 | ns |

*Note.* $\chi^2$ presents Kruskal-Wallis chi-squared statistics, and *Z* presents Dunn test z-statistics; RFDS: rise-fall difference score; ASD: autism spectrum disorder; SPC: sensory processing concern; TD: typical development; NA = not applicable; ns = not significant, \* $p < .05$ , \*\* $p < .01$ , \*\*\* $p < .001$ ; Bold typeface indicates effects that reached significance at the .05 level. P-values from pairwise comparisons were corrected for three comparisons using the Holm method.

## Discussion

### Altered Looming Bias in ASD

Our results provide electrophysiological evidence of auditory looming bias, the prioritized processing and perception of approaching auditory stimuli, in typically developing (TD) children and children with sensory processing concerns (SPC). Responses in these groups were characterized by a greater P1 peak amplitude during the rise phase than the fall phase of the stimuli used in the current study. In contrast, this finding was not present in the autism spectrum disorder (ASD) group. Furthermore, compared with the TD group, the ASD group had significantly lower Rise-Fall Difference Score (RFDS), meaning the asymmetry between the neural processing of looming and receding sounds is smaller in the ASD group.

To contextualize and interpret these group differences, we considered the cortical mechanism of looming bias put forward by Bidelman & Myers (19) and Ignatiadis et al.(46). Despite using different acoustic cues to induce looming and receding percepts (intensity-based in (19); spectrally defined in (46)), both studies’ connectivity analyses of EEG data revealed stronger information flow from the prefrontal cortex to the auditory cortex during looming than receding sounds, suggesting that looming bias may have a top-down neural mechanism. This suggests that our observed greater P1 peak amplitude during looming than receding phase of the stimuli reported in the TD and SPC groups in the current study may index a top-down effect from the anticipatory prefrontal cortex to auditory cortex; notably, this looming – receding difference is not present to the same extent in the ASD group.

Because looming bias has previously been suggested to reflect a perception asymmetry such that approaching signals are prioritized before receding and static ones (17), the ASD group’s altered looming bias observed in the current study may suggest an atypical ability to prioritize signals. In daily life, such an atypicality may lead to slower motor preparation upon detecting safety-relevant cues. For example, one may react more slowly to the sound of an approaching car engine or a rolling heavy object. Delayed motor readiness in these situations could reduce the time available for protective responses. Future studies should examine how alterations in looming bias in ASD are associated with changes in motor readiness.

Additionally, the ASD group’s reduced neural differentiation between looming and receding stimuli suggested altered context-dependent modulation of sensory input, a mechanism which may help to explain the Sensory Paradox (3). Specifically, the diminished phase-dependent weighting of sensory signals in the ASD group observed in the current study may point toward atypical dynamic adjustment of neural responses as contextual features change. This disruption in contextual modulation may help explain why sensory responsivity in ASD varies based on the context of a stimulus.

### Limitations

One limitation of the current study is that while stimuli were presented close in time (as often occurs with real-world looming stimuli), this rapid presentation rate precludes analysis of later ERP components, such as the P3 component. Future studies might extend stimulus length or include an inter-stimulus interval so that consecutive stimulus onsets are at least 600 ms apart, allowing these later components to be examined. Yet, because the speed and temporal structure of the tone cycle can influence neural responses, it will be critical to assess how the P1 component findings observed in the current design manifest under the conditions of the proposed new paradigm, particularly given prior evidence that sound intensity modulates P1 amplitude when stimuli are presented in rapid succession (38). The P3 component – often analyzed as two subcomponents of P3a and P3b – could be a focus of future investigation because prior work has pointed to P3’s association with neural processes closely related to looming bias, including stimuli change detection, attentional resource allocation, and context updating (47).

Future studies could also randomize the order of looming and receding stimuli. In the current study, looming stimuli always preceded receding stimuli. So, while we preserved ecological validity because looming stimuli are typically followed by receding stimuli (e.g., an approaching and departing car engine), randomizing looming and receding stimuli may offer the benefit of studying looming bias independent of other contextual factors such as the effect of habituation and novelty.

### Conclusion

Our findings point to the presence of an electrophysiological marker of altered auditory looming bias in ASD, indicating lack of prioritization of looming cues over receding ones. Additionally, while the current study did not detect significant differences in looming bias between the SPC group and either of the other two groups, this does not constitute evidence for the absence of a difference. Further research should continue to include the SPC group to evaluate the justification for, or against, establishing SPC as a standalone diagnosis. Overall, the findings suggest atypical use of temporal context to modulate auditory responses in young children with autism.

## Acknowledgments

The authors would like to thank the lab members who contributed to data collection and all the participants and their families for their valuable time and contributions.

## Data Availability Statement

Data are available upon reasonable request from the corresponding author.

## Ethics approval statement

The Institutional Review Board at Boston Children’s Hospital gave ethical approval for this work.

